# Combining Token Classification With Large Language Model Revision for Age-Friendly 4M Entity Recognition From Nursing Home Text Messages: Development and Evaluation Study

**DOI:** 10.64898/2026.03.31.26349861

**Authors:** Philip Amewudah, Mihail Popescu, Matthew S. Farmer, Kimberly R. Powell

## Abstract

**Background:** Secure text messages (TMs) exchanged among interdisciplinary care teams in nursing homes (NHs) contain clinical information that aligns with the Age-Friendly Health Systems 4Ms: What **M**atters, **M**edication, **M**entation, and **M**obility, yet, this information is not captured in any structured form, making it unavailable for systematic monitoring or quality reporting. Automatically extracting 4M information accurately and efficiently from these messages could enable several downstream applications within long term care settings. This task, however, is challenging because of the fragmented syntax, brevity, abbreviations, and informality of TMs.

**Objective:** This study aimed to develop and evaluate a multi-stage 4M Entity Recognition (4M-ER) pipeline that combines a fine-tuned token classifier with large language model (LLM) revision, using only locally deployed open-source models, to improve 4M information extraction from clinical TMs.

**Methods:** We used an expert-annotated dataset of 1,169 TMs representing conversations between interdisciplinary care teams across 16 Midwest NHs. The pipeline first identifies candidate text spans using a fine-tuned Bio-ClinicalBERT token classifier. A semantic similarity retriever then selects in-context exemplars to guide an LLM revision in which the LLM (Gemma, Phi, Qwen, or Mistral) performs boundary correction, label evaluation, and selective acceptance or rejection of candidate spans. Baselines for comparison included single-stage zero-shot LLMs, single-stage fine-tuned Bio-ClinicalBERT, and a fine-tuned LLM (Gemma) from a prior study. Ablation studies assessed the contribution of each pipeline stage and the effect of message filtering. Robustness was evaluated across 5 repeated runs.

**Results:** The 4M-ER pipeline outperformed the previously fine-tuned Gemma LLM across all 4M domains, achieving F_1_ (entity type) improvements of +2 to +11 percentage points without any additional fine-tuning and at roughly half the GPU memory (12 vs 24 GB). It also improved upon single-stage fine-tuned Bio-ClinicalBERT in Mobility, Mentation, and What Matters (+0.02 to +0.05 F_1_). Error analysis showed that LLM revision reduced false positives by 25% to 35% by correcting misclassifications caused by conversational ambiguity, while the fine-tuned Bio-ClinicalBERT’s high recall captured subtle entities that the fine-tuned Gemma missed. Silver data augmentation further improved the hardest domains, raising What Matters F_1_ from 0.59 to 0.67 and Mobility from 0.64 to 0.67. Ablation studies confirmed that restricting LLMs to revision only yielded optimal accuracy and efficiency.

**Conclusions:** The 4M-ER pipeline enables accurate and scalable extraction of 4M entities from clinical TMs by combining fine-tuned Bio-ClinicalBERT with LLM revision using only locally deployed open-source models. The structured 4M data produced by the pipeline can support 4M taxonomy and ontology construction, as demonstrated in the prior work, and provides a foundation for downstream applications including real-time clinical surveillance, compliance with emerging age-friendly quality measures, and predictive modeling in long-term care settings.

## Introduction

Nursing home care teams generate a continuous stream of clinically relevant information through secure text messages (TMs), yet almost none of it is captured in a form that can be monitored, aggregated, or reported, to the best of our knowledge. These TMs, exchanged through encrypted mobile platforms that support asynchronous coordination among nurses, physicians, therapists, and other staff [1,2], contain real-time observations about symptom changes, medication issues, cognitive status, mobility concerns, and resident or family preferences [3]. Much of this content maps directly to the 4M Age-Friendly Health Systems (AFHS) framework, which organizes care around 4 domains: What **M**atters, **M**edication, **M**entation, and **M**obility [4,5]. The 4M framework has been widely adopted in nursing homes (NHs) and other long-term care settings to guide clinical decision-making and quality improvement [6,7].

The problem is not that this 4M information is absent from care team communication; it is that the information is scattered across large volumes of conversational text and disappears after being read by the immediate recipient. No current system aggregates a resident’s 4M information across messages or makes this information available for compliance reporting under emerging quality measures such as the Centers for Medicare and Medicaid Services (CMS) Age-Friendly Hospital Measure. Automated extraction [8,9] could transform these informal communications into structured data that supports real-time clinical surveillance, care coordination and other downstream task like 4M taxonomy and ontology construction, as demonstrated in prior work [10]. However, the brevity, informality, and fragmented syntax of TMs make extraction challenging, requiring advanced Natural Language Processing (NLP) techniques [11].

NLP methods for clinical information extraction have advanced substantially in recent years. Named Entity Recognition (NER), the task of identifying and classifying clinical concepts in text, has become a foundational technique [12,13], and transformer-based architectures, particularly Bidirectional Encoder Representations from Transformers (BERT) and its domain-specific variants like Bio-ClinicalBERT, have achieved state-of-the-art NER performance on medical corpora [14–16]. More recently, large language models (LLMs) have shown significant promise for clinical extraction tasks [17].

Agrawal et al [18] demonstrated that few-shot LLM prompting can extract spans and relations from clinical notes, Hu et al [19] found that open-source LLMs achieve accuracy comparable to transformer encoders with greater contextual reasoning, and Guevara et al [20] used an LLM to identify social determinants of health from electronic health record narratives. A particularly relevant line of work has explored using LLMs not as standalone extractors but as correctors that refine encoder outputs. Sivarajkumar et al [21] proposed a 2-stage pipeline in which an LLM corrector applies domain rules to finalize cardiovascular event extractions, and Sreenivas et al [22] showed that coupling a retrieval-augmented LLM with a fine-tuned BERT extractor improved clinical NER by correcting span boundaries and resolving ambiguous entities. These studies establish the feasibility of encoder-LLM pipelines for clinical extraction, but none have applied this approach to the 4M framework.

Regarding 4M-specific extraction, most studies have targeted individual domains rather than all 4Ms collectively: medication change events from clinical notes [15], cognitive concerns and delirium indicators [23,24], and individual Mobility and What Matters concepts [25,26]. While the prior work by Farmer et al [10] and Powell et al [27] explored joint identification of 4M entities from clinical TMs, their approaches face limitations related to the high computational cost of LLM fine-tuning and performance variability with traditional classification methods [28]. No other existing approach jointly extracts all 4Ms from conversational TMs using a more computationally efficient framework.

To address this gap, this study introduces the 4M Entity Recognition (4M-ER) pipeline, a multi-stage framework designed to extract all 4M entities from clinical TMs accurately and efficiently. The key idea is to pair a high-recall encoder with LLM-based revision: the encoder identifies candidate entities from each message, a semantic retriever selects in-context exemplars from training annotations to guide the LLM, and the LLM reviews, refines, and finalizes the extracted entities. A message filtering strategy passes only TMs with candidate spans to the LLM, reducing computational overhead. We evaluate the 4M-ER pipeline against 3 baselines (zero-shot LLMs, standalone fine-tuned Bio-ClinicalBERT, and a fine-tuned Gemma LLM from the prior work by Farmer et al [10]), conduct ablation studies to isolate the contribution of each pipeline stage, and assess computational efficiency and robustness across repeated runs. The goal is a framework that can convert the 4M information already flowing through NH communication channels into structured data suitable for clinical surveillance, quality reporting, data standardization and predictive modeling.

## Methods

### Dataset and Annotation

From a corpus of TMs exchanged through a HIPAA-compliant communication platform [1] used for resident care coordination among interdisciplinary NH teams, 1,169 messages were selected and annotated as the ground truth for this study under a University of Missouri IRB-approved protocol. These are the same TMs used in the prior study on automatic 4M taxonomy development [10], reused here with the same annotation protocol for pipeline development and evaluation. Messages averaged approximately 17 words (SD 22.5) and reflected typical real-time clinical coordination in long-term care. For algorithm development, the ground truth dataset consisted of ∼75% for training and validation and ∼25% for testing (Table 1). Out of the training-validation set, ∼15% was set aside for validation in this study.

**Table 1.** Distribution of the ground truth dataset. Values represent the number (%) of messages containing at least 1 entity of the specified type.

|  | Training-Validation<br>dataset (n=860) | Testing dataset<br>(n=309) |
| --- | --- | --- |
| Medication | 144 (16.7%) | 97 (31.4%) |
| Mentation | 48 (5.6%) | 27 (8.7%) |
| Mobility | 55 (6.4%) | 30 (9.7%) |
| What Matters | 147 (17.1%) | 65 (21.0%) |
| At least one 4M | 290 (33.7%) | 162 (52.4%) |

A 4M annotation scheme aligned with the AFHS definitions was used to create the ground truth. Each TM was annotated by 2 clinical researchers with expertise in gerontology and nursing informatics for 1 or more of the following entity types:

- What Matters: references to patient or family goals, preferences, or care priorities.
- Medication: mentions of specific drugs, doses, routes, or administration concerns.
- Mentation: indicators of mood, cognition, delirium, or mental status.
- Mobility: descriptions of movement, transfers, falls, or ambulation capability.

Each message could contain multiple entities across 4M domains. For example, the TM *“Still in bed with periods of restlessness. Irregular heartbeat continues. Responding to verbal and verbal stimuli at this time”* was annotated with the entities (“*responding*”, Mentation), (“*verbal stimuli*”, Mentation), (“*restlessness*”, Mobility), and (“*in bed*”, Mobility). After independent annotation in the training-validation dataset, discrepancies were reviewed collaboratively and resolved through discussion. For instance, in the message *“She is a DNR. She has DX of HTN and other symbolic dysfunction,”* Annotator A labeled “*symbolic dysfunction*” as Mentation whereas Annotator B did not; following discussion, both agreed to include it under Mentation. The test dataset was subsequently annotated collaboratively.

Prior to resolving annotation disagreements in the training-validation set, Cohen K was computed for each 4M domain. Disagreements occurred in 28 Medication, 25 Mentation, 40 Mobility, and 93 What Matters instances, yielding almost perfect agreement for Medication (K = 0.89) and substantial agreement for Mentation (K = 0.73), Mobility (K = 0.61), and What Matters (K = 0.62), based on the Landis and Koch interpretive scale.

In the training-validation set, 16.7% of TMs contained Medication mentions, 17.1% contained What Matters, 5.6% contained Mentation, and 6.4% contained Mobility; 33.7% of training-validation messages contained at least one 4M mention or entity, while the test set had a higher 4M concentration (52.4%). Before processing, TMs were cleaned by removing nonstandard elements such as emojis and unknown characters. Stop words were retained to preserve the semantic meaning of extracted information spans.

### The 4M-ER Pipeline

The 4M-ER pipeline comprises 3 sequential stages: (1) a fine-tuned Bio-ClinicalBERT model performs token-level classification to identify candidate 4M entities, (2) a semantic retriever selects in-context exemplars from the ground truth annotations, and (3) an LLM reviser reviews and refines the candidate entities using these exemplars to produce the final 4M entities. An overview is shown in Figure 1.

**Figure 1.**
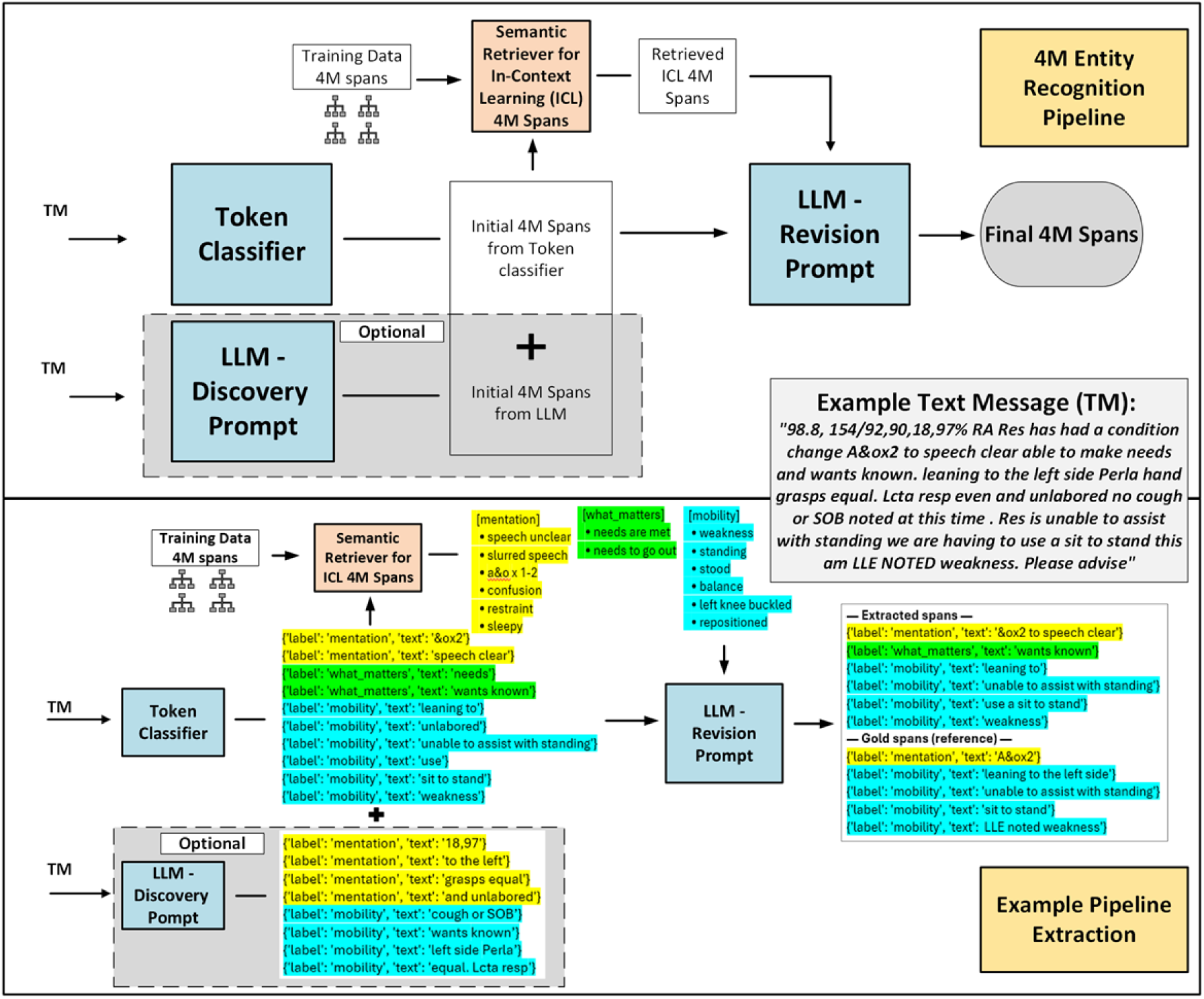
Overview of the 4M-ER pipeline. The system integrates (1) a fine-tuned token classifier for high-recall candidate span identification, (2) a semantic retriever that selects in-context exemplar spans from training annotations, and (3) an LLM revision prompt that performs boundary correction, label evaluation, and final acceptance or rejection of spans. The example message (right) shows how initial spans from the fine-tuned Bio-ClinicalBERT are refined into final 4M entities after LLM revision. TM: text message; ICL: in-context learning.

#### Stage 1: Candidate Entity Extraction With Fine-tuned Bio-ClinicalBERT

The first stage used Bio-ClinicalBERT [29,30], a bidirectional transformer pretrained on biomedical and clinical corpora, fine-tuned on the ground truth training dataset for 4M entity recognition. Input messages were tokenized with WordPiece encoding, and each token was labeled using the BIO (Begin, Inside, Outside) tagging scheme: B-XXX marks the beginning of a 4M entity, I-XXX its continuation, and O tokens outside any entity, where XXX denotes the 4M category (Medication, Mentation, Mobility, or What Matters). Because WordPiece splits some words into subword tokens (eg, “*restlessness*” becomes [‘*restless*’, *‘##ness’*]), subword tokens were aligned with their parent word’s BIO label to ensure consistent annotation. Fine-tuning hyperparameters included a learning rate of 1 × 10⁻⁵, batch size of 2, and up to 20 epochs, with the best model selected based on recall performance on the validation dataset.

During inference, only messages containing at least 1 predicted 4M entity were passed forward for LLM revision. This filtering strategy reduced the number of messages requiring LLM review, as most TMs do not contain any 4M entity (Table 1).

#### Stage 2: In-Context Exemplar Retrieval

After the fine-tuned Bio-ClinicalBERT proposes candidate spans, a semantic retriever links each candidate to exemplar spans drawn from the training annotations. Candidates and exemplars are embedded using the same sentence-level encoder (all-MiniLM-L6-v2 [31]), and cosine similarity is computed to rank exemplars within each candidate’s 4M label set. Each candidate is embedded as the text span plus a short left and right context window. Exemplars are stored as minimal tuples (label, span text, start, end). For each candidate, the retriever returns the top 3 exemplars ranked by similarity, with exact-match duplicates removed.

**Textbox 1.**
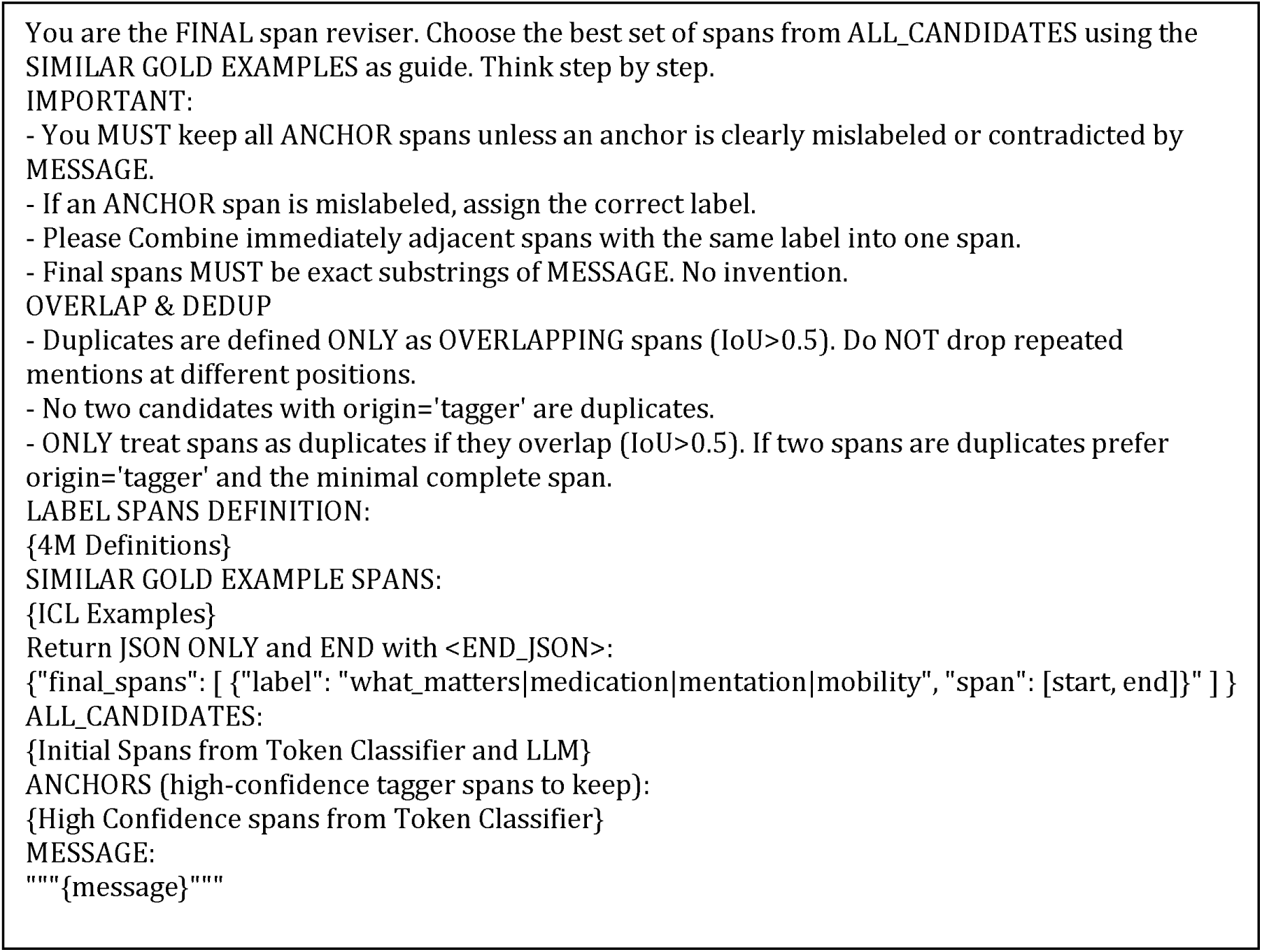
LLM revision prompt for the 4M-ER pipeline.

Retrieved items are formatted as span-text-and-label pairs and injected into the revision prompt alongside the candidate list.

#### Stage 3: LLM Revision

In the final stage, decoder-based LLMs contextualize and refine the candidate spans. 4 instruction-tuned models deployable on local GPUs were evaluated: Gemma-2-9B-IT (approximately 9 billion parameters) [32], Phi-3-Medium-4K-Instruct (approximately 14 billion parameters) [33], Qwen-2.5-14B-Instruct (approximately 14 billion parameters) [34,35], and Mistral-Nemo-Instruct-2407 (approximately 12 billion parameters) [36]. Each model received the original TM, the candidate spans from the fine-tuned Bio-ClinicalBERT, and the retrieved in-context exemplars.

A structured prompt (Textbox 1) instructed the LLM to review all candidate spans and determine the final set of entities. High-confidence spans (those with an average token-level classification probability from the fine-tuned Bio-ClinicalBERT > 0.9) were retained unless clearly mislabeled or contradicted by the message context. Mislabeled spans were corrected rather than discarded, adjacent same-label spans were merged, and duplicates were removed. The LLM output a JSON record containing entity type, text span, start and end indices, and confidence score.

All LLMs were run locally using 4-bit quantization to conserve GPU memory. Greedy decoding (selecting the single most probable token at each generation step) was used for all revision steps to ensure deterministic, schema-conformant outputs. Stochastic sampling methods (eg, temperature scaling, top-p sampling) were avoided to minimize output variance and hallucinated entity boundaries. Due to the clinical content of the data, only locally deployable open-source models were permitted by our Institutional Review Board (see Ethical Considerations).

### Baselines

3 baselines were used for comparison: (1) single-stage zero-shot prompting of each LLM for 4M extraction, using the prompt in Textbox A1 (Multimedia Appendix 1); (2) single-stage fine-tuned Bio-ClinicalBERT without LLM revision; and (3) the fine-tuned Gemma model used as a single-stage generative extractor from the prior work by Farmer et al [10].

### Ablation Studies

Ablation studies assessed the contribution of each pipeline component by varying (1) the role of the LLM (initial span generation vs revision only), (2) inclusion or exclusion of an optional zero-shot LLM discovery step, and (3) the effect of message filtering based on the fine-tuned Bio-ClinicalBERT outputs. In the discovery step, the LLM could propose additional candidate spans beyond those identified by the fine-tuned Bio-ClinicalBERT using the zero-shot prompt in Textbox A1 (Multimedia Appendix 1).

### Silver Data Augmentation

To evaluate whether the 4M-ER pipeline’s performance could be improved with additional training data, we developed a silver labeling pipeline to generate weakly labeled examples from unlabeled TMs. The pipeline operates in 3 stages: (1) candidate extraction, where an LLM extractor (gpt-oss:20b, run locally via Ollama [37]), a deterministic lexical matcher, and the gold-trained fine-tuned Bio-ClinicalBERT model independently propose candidate 4M spans; (2) adjudication, where the LLM classifies each candidate span as a valid 4M entity or not with a confidence score and evidence quote; and (3) skeptic review, where the LLM pass attempts to finally reject ambiguous labels. Only spans that maintained high confidence through both adjudication (2: 0.75) and skeptic review (2: 0.80) were retained, prioritizing precision over recall in the silver labels. This process produced additional 2,502 labeled messages from the TMs corpus, each filtered to contain at least 1 annotated entity.

Analysis of the silver dataset revealed a strong skew toward Medication entities (72.6% of entity tokens vs 61.7% in the gold data), with underrepresentation of Mentation, Mobility, and What Matters. To address this imbalance, we further filtered the silver dataset to retain only TMs where Medication entities comprised less than 50% of entity tokens. This reduced the silver training set from 2,502 to 636 examples, enriching the remaining data for underrepresented domains. The pre-trained Bio-ClinicalBERT was then fine-tuned using a 2-phase curriculum approach: Phase 1 fine-tuned the model on the filtered silver data for 5 epochs (learning rate 2 × 10⁻⁵), and Phase 2 continued fine-tuning from the best Phase 1 checkpoint under the same fine-tuning configuration used for the fine-tuned Bio-ClinicalBERT in the stage 1 of the 4M-ER pipeline. Full details of the silver labeling pipeline and distribution analysis are provided in Multimedia Appendix 1 and the 3 prompts used for candidate extraction, adjudication, and skeptic review are provided in Textboxes A2, A3, and A4 respectively.

### Evaluation Metrics

Performance was evaluated using the entity-type (ent_type) metric for NER-related tasks [38]. This metric evaluates whether the predicted label matches the ground truth category while tolerating small variations in span boundaries, which are common in short, conversational text where precise boundary agreement is often impractical. A prediction is considered correct if the label matches the ground truth, regardless of whether the span boundaries are exact. For example, the predicted Mobility entity “wheelchair” is considered correct relative to the ground truth “uses wheelchair” (same label, overlapping span), whereas “going to church” predicted as What Matters is incorrect relative to “attending” (semantically different span). A detailed comparison of ent_type against strict, exact, and partial match metrics is provided in Table A1 (Multimedia Appendix 1). F_1_ scores (harmonic mean of precision and recall) for ent_type matches were computed and used as the major metric for evaluation.

### Ethical Considerations

This study was conducted under a University of Missouri IRB-approved protocol. The dataset consisted of deidentified secure TMs collected through a HIPAA-compliant communication platform. Due to the clinical content of the data, the IRB restricted the use of closed-source commercial models (eg, ChatGPT [39] or Claude [40]); accordingly, only locally deployable open-source models were used for all experiments. No direct patient contact or intervention was involved, and all analyses were performed on previously collected and deidentified data.

### Implementation Environment

All experiments were run on a workstation with an NVIDIA RTX 4090 GPU (24 GB VRAM), Intel i9 processor, and 128 GB RAM. The key software dependencies were Python (version 3.10.18), Torch (version 2.4.1), Transformers (version 4.56.2) for Bio-ClinicalBERT fine-tunings and LLM deployment of the 4M-ER pipeline, Nervaluate (version 0.2.0) for evaluation and Ollama (version 0.18.1) for LLM calling during the silver label creation. Runtime and memory efficiency were measured for each LLM configuration on the test set (6533 word tokens; mean 21.1 tokens per message). A random seed of 66 was fixed for reproducibility.

## Results

### What the Pipeline Produces: A Clinical Example

Before presenting quantitative results, we illustrate what the 4M-ER pipeline produces on a real TM from the test set:

*“98.8, 154/92, 90, 18, 97% RA Res has had a condition change A&ox2 to speech clear able to make needs and wants known. leaning to the left side Perla hand grasps equal. Lcta resp even and unlabored no cough or SOB noted at this time. Res is unable to assist with standing we are having to use a sit to stand this am LLE NOTED weakness. Please advise”*

The 4M-ER pipeline extracted the following entities: (“*A&ox2 to speech clear*”, Mentation), (“*wants known*”, What Matters), (“*leaning to*”, Mobility), (“*unable to assist with standing*”, Mobility), (“*use a sit to stand*”, Mobility), (“*weakness*”, Mobility). The gold standard annotations for this message were: (“*A&Ox2*”, Mentation), (“*leaning to the left side*”, Mobility), (“*unable to assist with standing*”, Mobility), (“*sit to stand*”, Mobility), (“*LLE noted weakness*”, Mobility). The pipeline correctly identified the mentation assessment, the mobility concerns, and the patient’s expressed preferences. It also shows realistic boundary differences: “*leaning to*” rather than “*leaning to the left side,*” and “*weakness*” as a separate span rather than “*LLE noted weakness*.” These partial matches are scored as correct under the ent_type metric (see Methods), because the 4M category is right even when the span boundary is approximate.

### Extraction Performance

Table 2 summarizes precision, recall, and F_1_ (ent_type) across all configurations and 4M domains based on performances on the test dataset. The table reveals 3 key patterns. First, zero-shot LLMs failed across the board, averaging only 0.23 F_1_. Without domain-specific training, these models frequently produced overgeneralized outputs or misinterpreted clinical abbreviations, confirming that general-purpose LLMs alone are insufficient for 4M entity recognition in conversational TMs.

**Table 2.** Extraction performance across 4M domains for all model configurations. Values are presented as precision / recall / F_1_ (ent_type). Bold indicates the best F_1_ per domain.

| Configuration | Medication | Mentation | Mobility | What Matters |
| --- | --- | --- | --- | --- |
| Fine-tuned Bio-ClinicalBERT | 0.77 / 0.83 / **0.80** | 0.62 / 0.70 / 0.66 | 0.63 / 0.55 / 0.59 | 0.46 / 0.69 / 0.55 |
| Gemma (Zero Shot) | 0.36 / 0.47 / 0.41 | 0.07 / 0.20 / 0.10 | 0.08 / 0.08 / 0.08 | 0.10 / 0.19 / 0.13 |
| Phi (Zero Shot) | 0.34 / 0.23 / 0.27 | 0.16 / 0.23 / 0.19 | 0.15 / 0.08 / 0.11 | 0.28 / 0.17 / 0.22 |
| Qwen (Zero Shot) | 0.44 / 0.34 / 0.39 | 0.15 / 0.18 / 0.16 | 0.19 / 0.14 / 0.16 | 0.11 / 0.21 / 0.14 |
| Mistral (Zero Shot) | 0.38 / 0.36 / 0.37 | 0.07 / 0.18 / 0.10 | 0.12 / 0.12 / 0.12 | 0.08 / 0.20 / 0.12 |
| 4M-ER +<sup>a</sup> Gemma | 0.80 / 0.80 / **0.80** | 0.69 / 0.68 / 0.68 | 0.76 / 0.51 / 0.61 | 0.53 / 0.67 / **0.59** |
| 4M-ER + Phi | 0.79 / 0.77 / 0.78 | 0.72 / 0.65 / 0.68 | 0.81 / 0.53 / **0.64** | 0.53 / 0.63 / 0.57 |
| 4M-ER + Qwen | 0.79 / 0.80 / 0.79 | 0.71 / 0.68 / **0.69** | 0.81 / 0.51 / 0.63 | 0.51 / 0.66 / 0.58 |
| 4M-ER + Mistral | 0.79 / 0.80 / 0.79 | 0.69 / 0.60 / 0.64 | 0.70 / 0.53 / 0.60 | 0.49 / 0.69 / 0.57 |
| Fine-tuned Gemma<sup>b</sup> [10] | 0.82 / 0.74 / 0.78 | 0.62 / 0.67 / 0.64 | 0.69 / 0.46 / 0.55 | 0.46 / 0.52 / 0.48 |

<sup>a</sup>4M-ER configurations use fine-tuned Bio-ClinicalBERT for candidate extraction followed by the named LLM for revision
<sup>b</sup>Fine-tuned Gemma is a single-stage generative extractor from prior work, not a 4M-ER configuration.

Second, LLM revision is primarily a precision mechanism (Table 2). The most dramatic gain was in Mobility, where precision increased from 0.63 (fine-tuned Bio-ClinicalBERT alone) to 0.81 (Phi and Qwen configurations). This means that when the pipeline flagged a Mobility entity, it was correct 81% of the time compared with 63% without revision. For Mentation, precision rose from 0.62 to 0.72 (Phi configuration). For What Matters, precision improved from 0.46 to 0.53 (Gemma configuration). In practical terms, these gains mean fewer false alerts if the pipeline were used in a clinical monitoring context. Recall remained stable across configurations, indicating that LLM revision improves the reliability of extracted entities without sacrificing the encoder’s ability to detect them. For Medication, precision was already relatively high (0.77) and improved to 0.80 (Gemma configuration), likely because medication entities (drug names, doses) have well-defined lexical boundaries that benefit less from contextual revision.

Third, the 4M-ER pipeline matched or exceeded the fine-tuned Gemma LLM from prior work [10] across all domains, without any additional model fine-tuning (Table 2). The fine-tuned Gemma required full model training on the same dataset; the 4M-ER pipeline used Gemma only at inference time. Despite this, the Gemma-based pipeline configuration alone achieved F_1_ of 0.80, 0.68, 0.61, and 0.59 compared with the fine-tuned Gemma’s 0.78, 0.64, 0.55, and 0.48, representing improvements of +2 to +11 percentage points across domains. The pipeline also showed higher recall than the fine-tuned Gemma in every domain, reflecting the advantage of the fine-tuned Bio-ClinicalBERT’s high-recall extraction as the first pipeline stage.

Finally, different LLM revisers showed domain-specific strengths. Gemma performed best for Medication (F_1_ 0.80, precision 0.80) and What Matters (F_1_ 0.59, precision 0.53), Qwen for Mentation (F_1_ 0.69, precision 0.71), and Phi for Mobility (F_1_ 0.64, precision 0.81). No single LLM reviser dominated across all 4 domains, suggesting that the choice of reviser interacts with the linguistic characteristics of each domain and that an adaptive routing strategy could yield further gains.

### Ablation Analysis

Ablation experiments tested which components of the pipeline contribute most to performance (Figure 2). Two findings stand out. First, restricting LLMs to revision only (rather than also generating initial spans) yielded the best balance of accuracy and efficiency. When LLMs were used for both span generation and revision, performance did not improve and sometimes degraded, suggesting that the encoder’s high-recall extraction is a better starting point than LLM-generated candidates. Second, filtering messages to pass only those with preliminary fine-tuned Bio-ClinicalBERT spans to the LLM improved both efficiency and accuracy: Mentation F_1_ increased from 0.62 to 0.64, and Mobility from 0.59 to 0.63. Together, these results confirm that the LLM functions optimally as a selective refinement stage applied to a high-recall encoder output, not as a general-purpose extractor.

**Figure 2.**
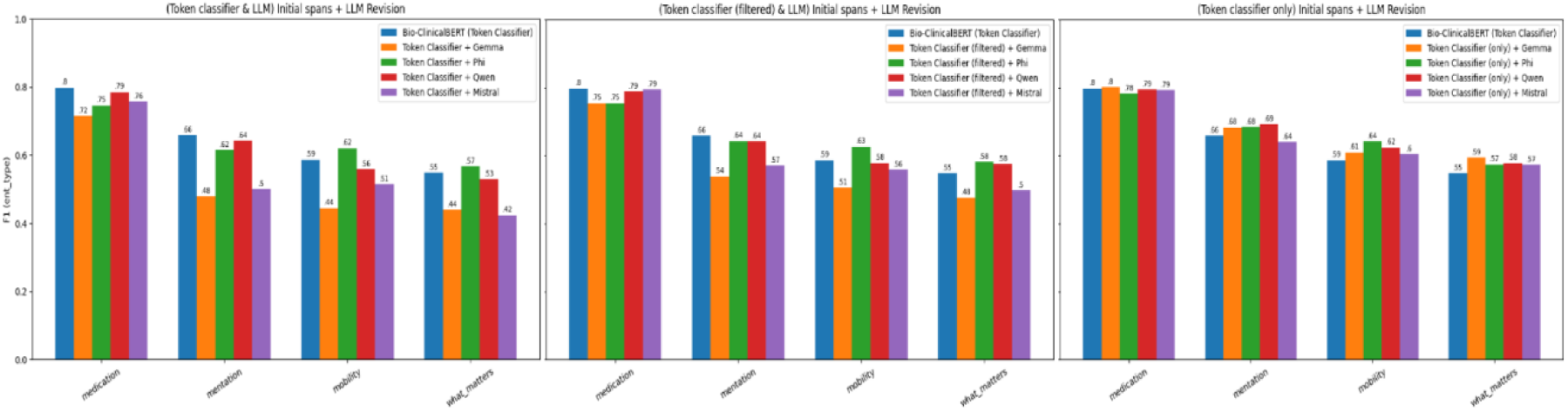
Ablation studies of varying LLM roles and message filtering strategies. The rightmost configuration (token classifier initial spans + LLM revision) is the main 4M-ER pipeline.

### Error Analysis

To understand what the 4M-ER pipeline gets right that the baselines miss, and where it still fails, we examined specific error patterns across model configurations (Tables 3, 4, and 5).

**Table 3.** False-positive counts by 4M domain across model configurations. The 4M-ER pipeline produces substantially fewer false positives than the standalone fine-tuned Bio-ClinicalBERT and the fine-tuned Gemma baseline.

| Configuration | Count of FP Spans |  |  |  |
| --- | --- | --- | --- | --- |
|  | Medication | Mentation | Mobility | What Matters |
| Fine-tuned Bio-ClinicalBERT | 38 | 9 | 16 | 69 |
| Fine-tuned Bio-ClinicalBERT + Gemma revision | 33 | 6 | 10 | 50 |
| Fine-tuned Bio-ClinicalBERT + Mistral revision | 31 | 6 | 12 | 61 |
| Fine-tuned Bio-ClinicalBERT + Phi revision | 31 | 5 | 9 | 46 |
| Fine-tuned Bio-ClinicalBERT + Qwen revision | 31 | 5 | 10 | 52 |
| Fine-tuned Gemma (LLM from prior study) | 21 | 12 | 13 | 53 |

FP: predicted entity with no matching ground truth entity of the same label

**Table 4.** False-negative counts by 4M domain across model configurations. The 4M-ER pipeline reduces false negatives relative to the fine-tuned Gemma model for nearly all labels.

| Configuration | Counts of FN Spans |  |  |  |
| --- | --- | --- | --- | --- |
|  | Medication | Mentation | Mobility | What Matters |
| Fine-tuned Bio-ClinicalBERT | 26 | 12 | 22 | 26 |
| Fine-tuned Bio-ClinicalBERT + Gemma revision | 30 | 13 | 24 | 28 |
| Fine-tuned Bio-ClinicalBERT + Mistral revision | 31 | 14 | 24 | 27 |
| Fine-tuned Bio-ClinicalBERT + Phi revision | 34 | 14 | 23 | 32 |
| Fine-tuned Bio-ClinicalBERT + Qwen revision | 30 | 13 | 24 | 29 |
| Fine-tuned Gemma (LLM from prior study) | 44 | 16 | 32 | 43 |

FN: ground truth entity with no matching prediction of the same label

**Table 5.** Sample predicted annotations and gold annotations. Pipeline annotations are from 4M-ER + Gemma revision configuration.

|  |  |  |
| --- | --- | --- |
| (1) | **Message** | Need #####<sup>c</sup> chart brought to DNS office |
|  | **Gold Annotation** | () |
|  | **Baseline Annotation <sup>a</sup>** | (*'what matters: DNS'*) |
|  | **Pipeline Annotation** | () |
| (2) | **Message** | Range of motion performed limited on LLE due to pain. Resident cries out in pain during assessment. Difficulty with mobility. Slightly bearing weight. Unsteady. Still complaining of pain tearful and facial grimacing. No PRN pain reliever order. Concerned about pain for this resident. Can we have a Tylenol order? Also would you like further x Ray testing to rule out the femur? |
|  | **Gold Annotation** | (*'what matters: pain', 'what matters: cries out in pain during assessment', 'what matters: complaining of pain', 'what matters: tearful', 'medication: Tylenol'*) |
|  | **Baseline Annotation <sup>a</sup>** | (*'mobility: Range of motion', 'what matters: pain', 'what matters: cries out in pain', 'mobility: mobility', 'mobility: bearing weight. Unsteady', 'what matters: complaining of pain tearful', 'mentation: facial grimacing', 'medication: PRN', 'what matters: pain', 'medication: reliever', 'what matters: pain', 'medication: Tylenol'*) |
|  | **Pipeline Annotation** | (*'what matters: pain', 'what matters: cries out in pain', 'mobility: mobility', 'mobility: bearing weight. Unsteady', 'what matters: complaining of pain tearful', 'what matters: pain', 'what matters: pain', 'medication: Tylenol'*) |
| (3) | **Message** | ##### saw pt. encourage to use stand up lift. pt refuses hoyer 2x assist if pt refuses stand up lift. pt and regular staff state pt does well w/ 2 person transfer. tiffany |
|  | **Gold Annotation** | (*'what matters: pt refuses hoyer', 'what matters: refuses', 'mobility: stand up lift', 'mobility: 2 person transfer'*) |
|  | **Baseline Annotation <sup>b</sup>** | (*'mobility: stand up lift', 'mobility: hoyer', 'mobility: 2 person transfer'*) |
|  | **Pipeline Annotation** | (*'what matters: pt', 'mobility: stand up lift', 'what matters: pt refuses', 'mobility: hoyer', 'what matters: pt refuses', 'mobility: stand up lift', 'what matters: regular', 'what matters: pt does well', 'mobility: 2 person transfer'*) |
| (4) | **Message** | Spoke with ##### with Dr ##### office. Dr gave orders to Dc her equalizer boot. Does not want a repeat X-ray. |
|  | **Gold Annotation** | (*'mobility: equalizer boot', 'mobility: X-ray'*) |
|  | **Baseline Annotation <sup>b</sup>** | () |
|  | **Pipeline Annotation** | (*'mobility: equalizer boot'*) |
| (5) | **Message** | pt family here on visit, noticed pt is not responding as normal, slurring in speech and really has no energy, vs taken, 98.6,88,18,131/85, family wants pt to be sent out to make sure that tumors have not effected him in any new way |
|  | **Gold Annotation** | (*'what matters: family here on visit', 'what matters: family wants pt to be sent out', 'mentation: not responding', 'mentation: slurring in speech', 'mentation: has no energy'*) |
|  | **Baseline Annotation <sup>b</sup>** | (*'what matters: family', 'what matters: wants pt to be sent out', 'mentation: not responding as normal', 'mentation: slurring in speech', 'mobility: no energy'*) |
|  | **Pipeline Annotation** | (*'what matters: family', 'mentation: pt is not responding as normal, slurring in speech', 'mentation: energy', 'what matters: pt to be sent out'*) |

<sup>a</sup>Annotation from the Single-stage fine-tuned Bio-ClinicalBERT without LLM evaluation
<sup>b</sup>Annotation from the fine-tuned Gemma model from prior study [10]
<sup>c</sup>##### represents name mentions in the TMs

#### False positives corrected by LLM revision

The LLM revision stage was most effective at correcting FPs caused by conversational ambiguity, where a token classifier assigns a 4M label based on lexical similarity without understanding the sentence-level context. What Matters had the highest FP rates across all models. In the message *“Need #### chart brought to DNS office,”* (where #### represents name mentions in the TMs) the fine-tuned Bio-ClinicalBERT misclassified “*DNS*” as What Matters, likely because the abbreviation resembles “*DNR*” (do not resuscitate), a concept central to the What Matters domain. The LLM revision stage, which processes the full message, recognized that “*DNS office*” refers to an administrative location and correctly rejected the span. Similarly, in *“…No PRN pain reliever order…,”* the fine-tuned Bio-ClinicalBERT flagged “*PRN*” and “*reliever*” as Medication because these tokens are semantically close to medication concepts. The LLM recognized from context that the message was describing the absence of an order, not a medication entity, and removed both spans (Table 5). Across all domains, the pipeline consistently reduced FP counts relative to the standalone fine-tuned Bio-ClinicalBERT (Table 3), with the largest reductions in What Matters and Medication.

#### False negatives reduced by high-recall encoding

The fine-tuned Gemma baseline had the highest FN rates across most domains, particularly for entities expressed through subtle or indirect clinical language. For instance, the fine-tuned Gemma missed “*pt refuses hoyer*” and “*refuses*” in *“…pt refuses hoyer 2x assist if pt refuses stand up lift…”* (What Matters) and “*equalizer boot*” in *“…Dr gave orders to Dc her equalizer boot…”* (Mobility). In both cases, the clinical meaning is implied rather than stated explicitly (Table 5, examples 3-4). The fine-tuned Bio-ClinicalBERT’s token-level training captured many of these entities that the fine-tuned Gemma missed, and the 4M-ER pipeline inherited this advantage: FN counts were reduced relative to the fine-tuned Gemma across all domains (Table 4).

#### Remaining errors: label-level misclassification

The most persistent error type was assigning the correct span boundaries but the wrong 4M category, particularly confusing Mentation with Mobility or What Matters (Figure A2, Multimedia Appendix 1). This confusion reflects genuine clinical ambiguity. In the message *“pt family here on visit, noticed pt is not responding as normal, slurring in speech and really has no energy…,”* the gold standard annotated “*has no energy*” as Mentation (a cognitive/mental status observation), but the fine-tuned Gemma labeled “*no energy*” as Mobility (Table 5, example 5). The 4M-ER pipeline correctly categorized it as Mentation but captured only “*energy*” rather than “*has no energy*.” Expressions like “*no energy,*” “*restlessness,*” and “*not responding*” sit at the boundary between Mentation and Mobility because they can reflect either a cognitive state or a physical limitation depending on context. The 4M-ER pipeline with Gemma revision reduced these misclassifications to the lowest levels across all configurations (4 Mentation-as-Mobility, 2 Mentation-as-What Matters, 1 Medication confusion), but did not eliminate them entirely. These remaining errors reflect the inherent overlap among 4M domains in conversational clinical language and represent a target for future work.

### Effect of Silver Data Augmentation

Fine-tuning Bio-ClinicalBERT on the combined gold and silver data using curriculum learning improved the 4M-ER pipeline’s performance, with the largest gains in the domains that were hardest to extract with gold data alone (Table 6). The silver-augmented fine-tuned Bio-ClinicalBERT achieved F_1_ of 0.78 (Medication), 0.69 (Mentation), 0.66 (Mobility), and 0.62 (What Matters), compared with 0.80, 0.66, 0.59, and 0.55 respectively for the gold-only encoder. Mobility and What Matters improved by +0.07 each, reflecting the benefit of exposing the model to a broader range of entity patterns for these linguistically diverse categories during silver pretraining. When the silver-augmented encoder was combined with LLM revision, the best configurations achieved F_1_ of 0.81 (Medication, Qwen), 0.73 (Mentation, Phi), 0.67 (Mobility, Qwen), and 0.67 (What Matters, Qwen). Compared with the best gold-only pipeline results, this represents additional gains of +0.01 (Medication), +0.04 (Mentation), +0.03 (Mobility), and +0.08 (What Matters). The What Matters improvement from 0.59 to 0.67 is particularly notable, as this domain had been the most challenging due to the conversational, context-dependent nature of patient preference expressions.

**Table 6.** Best 4M-ER pipeline performance with and without silver data augmentation. Values are presented as precision / recall / F_1_ (ent_type). Bold indicates the best F_1_ per domain.

| Configuration | Medication | Mentation | Mobility | What Matters |
| --- | --- | --- | --- | --- |
| Best gold-only 4M-ER | .80 / .80 / .80 | .71 / .68 / .69 | .81 / .53 / .64 | .53 / .67 / .59 |
| Best gold+silver 4M-ER <sup>a</sup> | .85 / .77 / **.81** | .73 / .73 / **.73** | .83 / .56 / **.67** | .70 / .64 / **.67** |

<sup>a</sup> Best LLM revisions – Qwen for What Matters, Medication and Mobility; Phi for Mentation

### Computational Efficiency and Robustness

The 4M-ER pipeline achieved better extraction performance than the fine-tuned Gemma model from the prior work [10] while using substantially fewer computational resources. Among the LLM revisers, Qwen was the fastest (26 minutes 3 seconds on the test set), followed by Gemma (38 minutes 6 seconds), Mistral (90 minutes 48 seconds), and Phi (107 minutes 54 seconds). Message filtering reduced the number of TMs processed by LLMs by approximately 35%. Nearly all 4M-ER configurations (except Phi) required less than 12 GB VRAM during inference, compared with more than 12 GB required for fine-tuning the Gemma model.

Robustness was assessed by running each configuration 5 independent times under same conditions. All configurations demonstrated high stability under greedy decoding, with run-to-run F_1_ (ent_type) variations typically within ±0.005 to ±0.02 across all domains (Figure A1, Multimedia Appendix 1). This level of reproducibility confirms that the pipeline produces consistent results without stochastic drift, supporting its reliability for clinical deployment.

## Discussion

### Principal Findings

The central finding of this study is that pairing a domain-specific encoder with inference-only LLM revision produces better 4M extraction than either approach alone, and does so more efficiently than fine-tuning an LLM end-to-end. The 4M-ER pipeline achieved F_1_ improvements of +2 to +11 percentage points over a fully fine-tuned Gemma LLM at half the GPU cost, without requiring any additional model training. The precision and recall breakdown reveals why: LLM revision is primarily a precision mechanism. For Mobility, precision jumped from 0.63 to 0.81 after revision, meaning that 4 out of 5 Mobility entities the pipeline flagged were correct. For Mentation, precision rose from 0.62 to 0.72. Recall, meanwhile, remained stable, confirming that fine-tuned Bio-ClinicalBERT’s high-recall extraction provides a reliable foundation that the LLM refines rather than replaces. The ablation results confirmed that this division of labor (encoder for extraction, LLM for revision) is the optimal configuration; using the LLM for both roles degraded rather than improved performance. Silver data augmentation further improved the pipeline’s performance ceiling, with the largest gains in the 2 most challenging domains: What Matters F_1_ increased from 0.59 to 0.67, and Mobility from 0.64 to 0.67.

### Comparison With Prior Work

Most prior work on 4M-related extraction has focused on individual domains using fine-tuned models applied to formal clinical narratives [15,24,25,25,26]. These approaches face 2 limitations when applied to TMs in long-term care: the terse, context-sparse nature of the text degrades model performance, and training a separate model per domain could be impractical for collective 4M extraction. The 2 studies that attempted joint 4M extraction from TMs [10,27] relied on either full LLM fine-tuning (computationally expensive) or traditional classification methods (limited accuracy). The 4M-ER pipeline addresses both issues. By using the fine-tuned Bio-ClinicalBERT as a shared encoder across all 4 domains and adding LLM revision as a lightweight reasoning layer, it achieves collective extraction without per-domain models and without the cost of LLM fine-tuning. The +2 to +11 F_1_ improvement over the fine-tuned Gemma model from Farmer et al [10], achieved at half the GPU memory, demonstrates that strategic encoder-decoder pairing can outperform brute-force fine-tuning for this task.

### Implications for Age-Friendly Health Systems

The most immediate benefit of the 4M-ER pipeline is enabling structured, real-time clinical surveillance from a data source that already exists but is currently invisible to systematic monitoring. NH care teams already communicate 4M information through TMs as part of routine care, but this information is read once by the recipient and never aggregated. When a resident’s TMs mention mobility concerns in the morning, cognitive changes at midday, and a family preference for comfort care in the afternoon, no current system connects those signals across shifts and team members to identify concurrent decline across multiple 4M domains. The 4M-ER pipeline can make this cross-shift, cross-domain aggregation possible by transforming each TM into structured 4M entities as it arrives.

A related near-term application is shift handoff support. Communication during shift changes in long-term care is often fragmented, and clinically relevant 4M information discussed during the preceding shift may not be fully conveyed to the incoming team. Structured 4M output from the pipeline could be used to auto-generate shift-level summaries of what changed across each domain for each resident, providing incoming staff with an organized view of the day’s 4M activity.

The pipeline also addresses a pressing compliance need. The 4M framework is now incorporated into the CMS Hospital Inpatient Quality Reporting program, and health systems will increasingly need to demonstrate 4M-aligned care delivery. In long-term care, much of the evidence that this care is happening lives in unstructured TMs that cannot be audited at scale. The 4M-ER pipeline turns this unstructured evidence into reportable, auditable data, supporting quality measurement and performance reporting aligned with the AFHS initiative.

Beyond these immediate applications, the structured 4M data generated by the pipeline can support longer-term goals including standardized ontology construction for 4M interoperability and predictive modeling of resident outcomes such as avoidable hospital transfers as demonstrated in the prior works by Farmer et al [10] and Powell et al [27] respectively.

### Methodological Contributions

This study makes methodological contributions to clinical NLP beyond the 4M application. First and foremost, it provides evidence that revision-oriented encoder-decoder architectures, where a lightweight LLM refines encoder predictions at inference time, can match or exceed the performance of fully fine-tuned LLMs while remaining accessible to institutions with limited GPU resources. This finding has implications for any clinical NER task where fine-tuning large models is impractical. Second, the message filtering strategy, which substantially reduced the LLM workload without sacrificing accuracy, demonstrates a scalable approach to managing computational cost in streaming data environments. Third, the silver data generation and augmentation approach, which combines LLM-based extraction with adversarial skeptic review, class-imbalance-aware filtering, and 2-phase curriculum training, offers a reusable strategy for expanding limited gold-annotated clinical datasets while controlling label noise. This approach is particularly relevant for clinical NLP tasks where expert annotation is expensive and the most clinically important entity categories are also the rarest in training data. Fourth, the study validates the use of the ent_type evaluation metric for clinical TMs, where entity boundaries are inherently imprecise and strict span matching would penalize clinically valid extractions.

### Limitations and Future Directions

The dataset, though drawn from 16 NHs, represents a single geographic region, which may limit generalizability to care teams with different communication styles or platforms. While LLM revision substantially improved precision across all domains, recall remained modest for some domains with gold-only training (eg, Mobility recall of 0.51-0.53).

Silver data augmentation partially addressed this gap, improving Mobility F_1_ from 0.64 to 0.67 and What Matters from 0.59 to 0.67, but further gains may require larger or more domain-balanced silver datasets. Future work should evaluate whether the current extraction accuracy is sufficient for specific clinical applications (eg, shift-level surveillance vs real-time alerting) and determine what level of clinician oversight is needed before deployment. Only medium-sized open-source LLMs and one pre-trained Bio-ClinicalBERT were evaluated; future work should also assess whether larger models or domain-specific instruction tuning further improves revision quality.

Next steps include extending the 4M-ER pipeline with ontology-driven span normalization using a 4M knowledge graph, prospective deployment within NH communication platforms for real-time 4M tracking, and evaluation of its effect on predicting adverse events such as avoidable hospital transfers. A particularly promising direction is an adaptive 4M-ER pipeline that dynamically routes each candidate span to the most effective LLM reviser on the basis of domain. Our results indicated domain-specific strengths (Gemma for Medication and What Matters, Qwen for Mentation, Phi for Mobility), suggesting that a domain-specialized ensemble system could further improve precision and recall while maintaining efficiency.

## Conclusions

The 4M-ER pipeline, combining a fine-tuned Bio-ClinicalBERT encoder with inference-only LLM revision, provides a scalable and resource-efficient approach for extracting Age-Friendly 4M entities from clinical TMs. The pipeline improved extraction accuracy across all 4M domains compared with standalone models and the previously fine-tuned Gemma LLM [10], while requiring fewer computational resources and maintaining high reproducibility. Silver data augmentation further improved performance in the most challenging domains, demonstrating that the pipeline’s accuracy can scale with additional training data. By transforming unstructured care team communications into structured 4M data, this pipeline can support 4M taxonomy and ontology construction, as demonstrated in the prior study [10], and provides a foundation for downstream applications including real-time clinical surveillance, compliance with emerging age-friendly quality measures, and predictive modeling in long-term care settings.

## Data Availability

Due to the presence of potentially identifiable clinical information and institutional data use agreements, the dataset cannot be publicly shared. Requests for data access will be reviewed on a case-by-case basis and require a signed data use agreement with the corresponding institution. All code used to develop and evaluate the 4M-ER pipeline is available on GitHub

## Acknowledgments

This work was supported by the National Institute on Aging of the National Institutes of Health under award R01AG078281. The content is solely the responsibility of the authors and does not necessarily represent the official views of the National Institutes of Health.

## Data and Codes Availability

Due to the presence of potentially identifiable clinical information and institutional data use agreements, the dataset cannot be publicly shared. Requests for data access will be reviewed on a case-by-case basis and require a signed data use agreement with the corresponding institution. All code used to develop and evaluate the 4M-ER pipeline is available on GitHub [41].

## Abbreviations

4M-ER: 4M Entity Recognition
AFHS: Age-Friendly Health Systems
BERT: Bidirectional Encoder Representations from Transformers
BIO: Begin, Inside, Outside
CMS: Centers for Medicare and Medicaid Services
FN: false negative
FP: false positive
GPU: graphics processing unit
HIPAA: Health Insurance Portability and Accountability Act
ICL: in-context learning
IRB: Institutional Review Board
LLM: large language model
NER: Named Entity Recognition
NH: nursing home
NLP: Natural Language Processing
TM: text message
VRAM: video random access memory

## Authors’ Contributions

PA: conceptualization, methodology, software, formal analysis, investigation, visualization, writing - original draft. KP: funding acquisition, supervision, writing - review and editing. MP: supervision, writing - review and editing. MF: writing - review and editing. All authors reviewed and approved the final manuscript.

## Multimedia Appendix 1

**Table A1.**
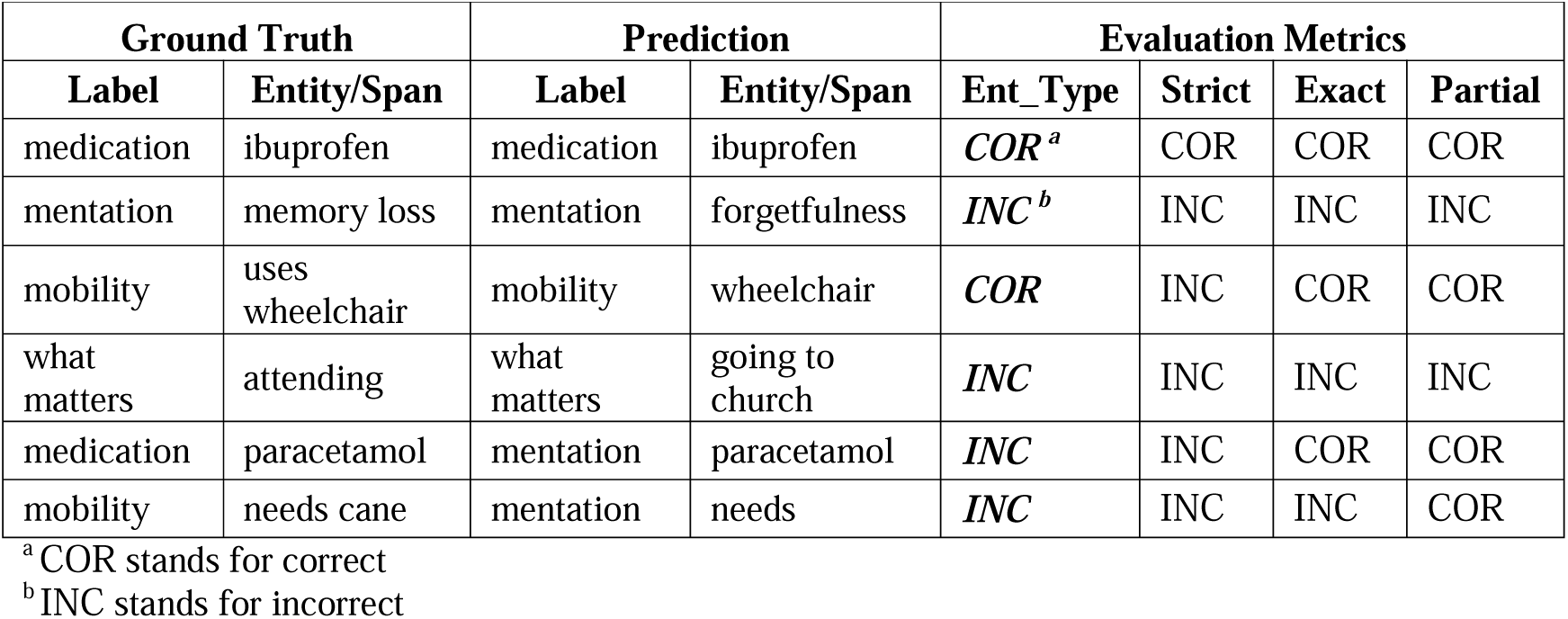
Comparison of evaluation metrics for NER-related tasks (ent_type, strict, exact, and partial match).

**Textbox A1.**
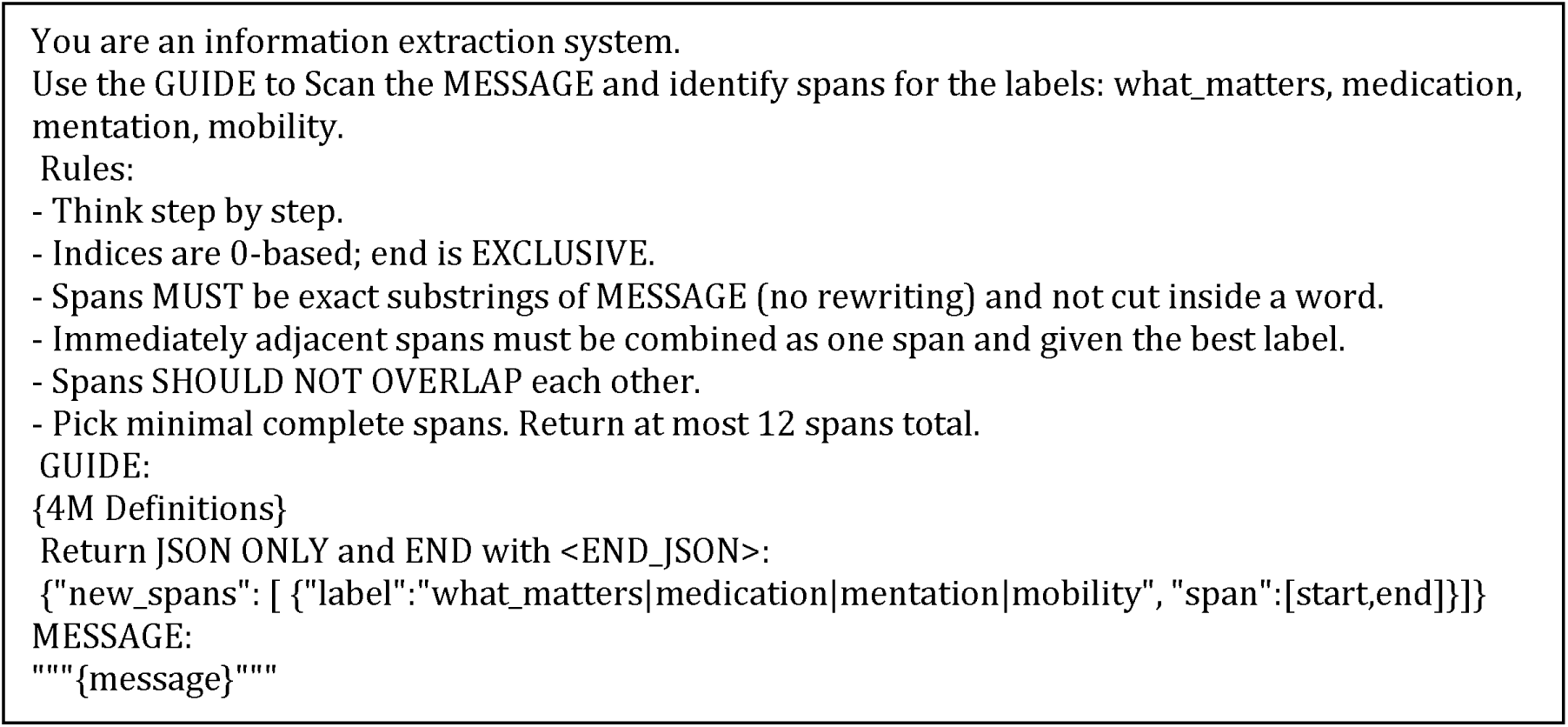
Zero-shot prompt for optional 4M candidate span detection by LLMs.

**Figure A1.**
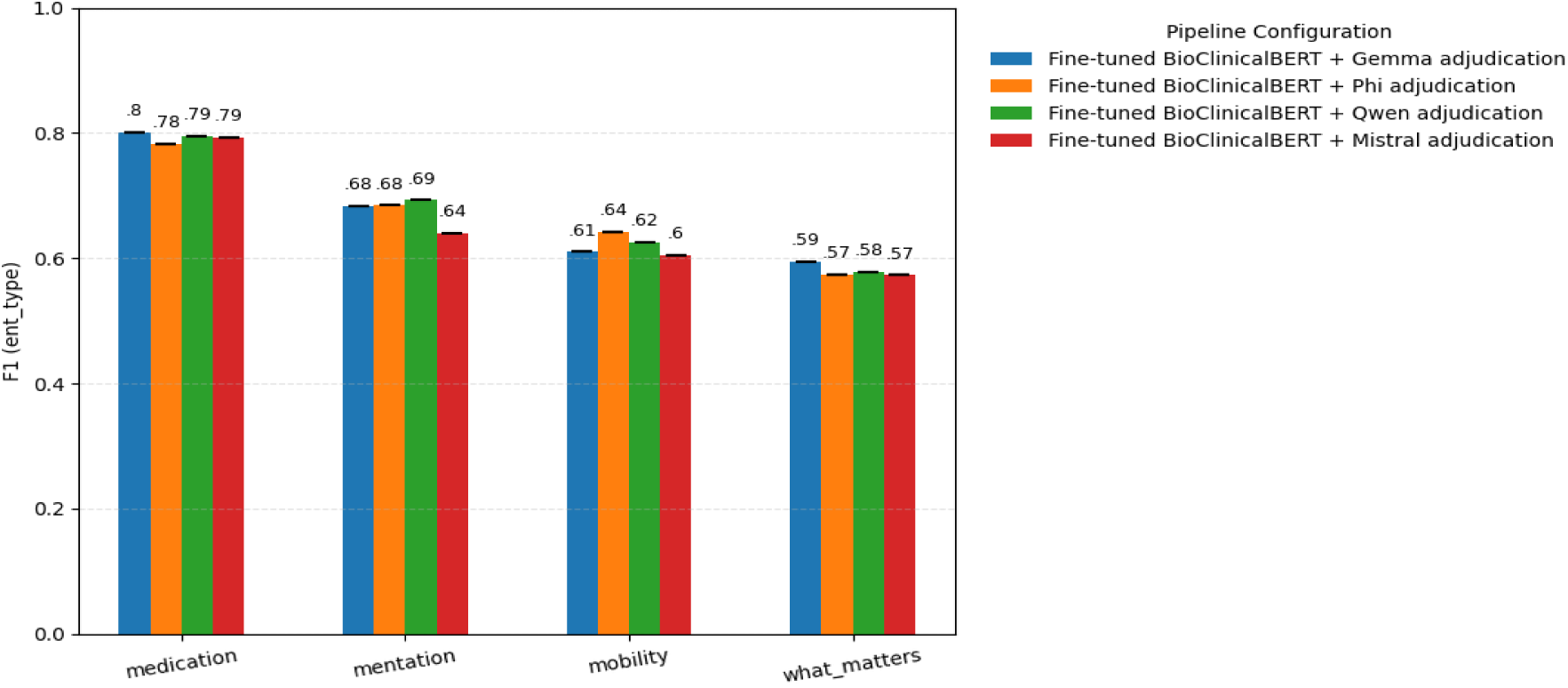
Robustness analysis of 4M-ER pipeline configurations. Mean F (ent_type) scores with SD whiskers are shown across 5 repeated runs for each 4M domain. All configurations exhibit minimal variance (typically ≤ 0.02), demonstrating high run-to-run stability.

**Figure A2.**
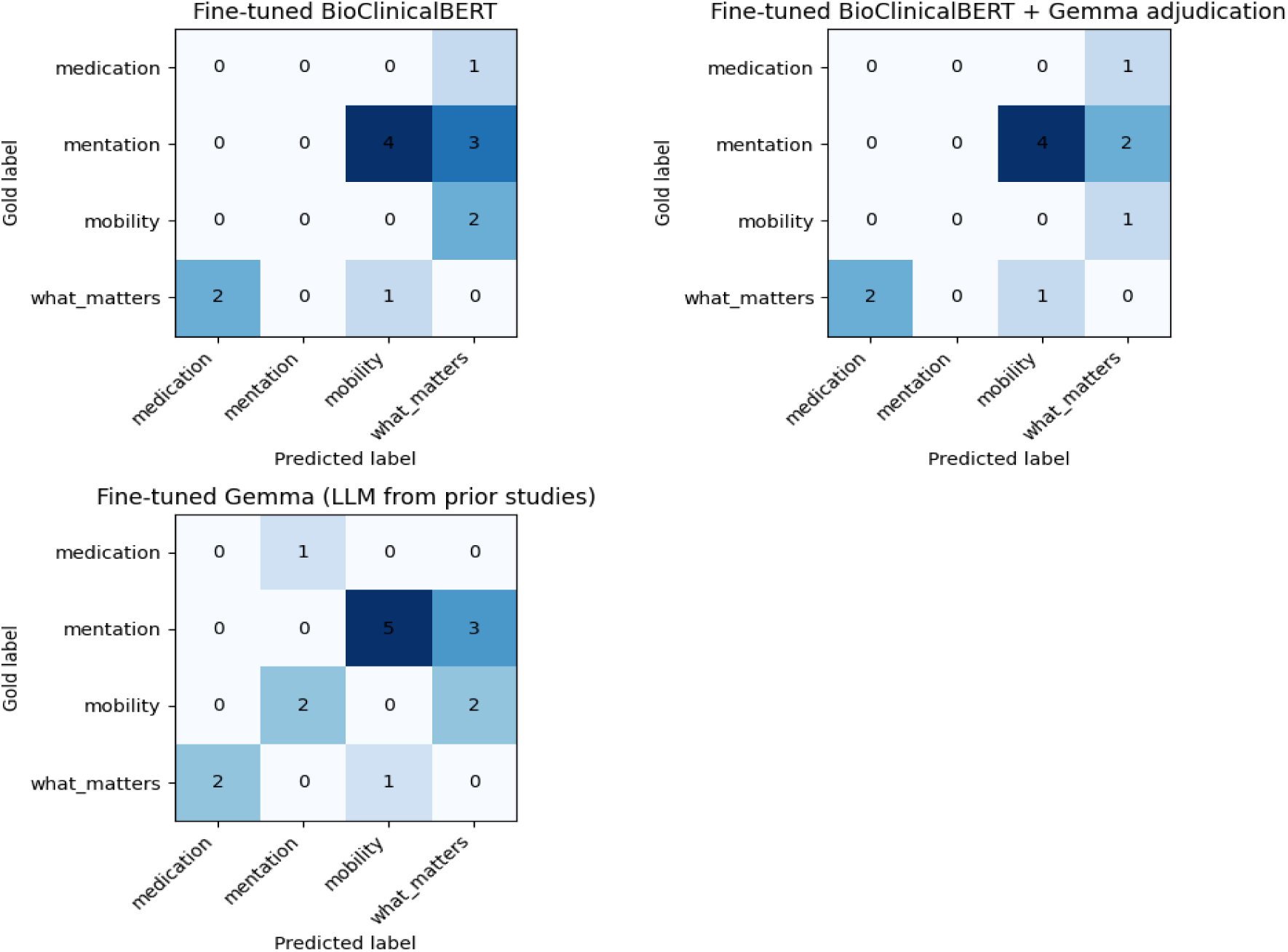
Confusion matrices comparing misclassification patterns for Bio-ClinicalBERT alone, Bio-ClinicalBERT + Gemma revision, and the fine-tuned Gemma model. LLM revision reduces cross-label confusion, particularly misclassification of Mentation as What Matters and Mobility as What Matters.

### Appendix Text

Silver Data Generation Pipeline. Full description of the 3-stage silver labeling pipeline (candidate extraction, adjudication, and skeptic review), data distribution analysis, and silver data filtering strategy.

#### Overview

To augment the limited gold-annotated training set we developed a multi-stage silver labeling pipeline that combines large language model reasoning with deterministic lexical matching to produce high-confidence weakly labeled training data. The pipeline was designed to prioritize precision over recall, under the premise that noisy false positives are more damaging to downstream model training than missed entities. All LLM calls were executed locally using gpt-oss:20b through the Ollama framework (v0.18.1).

#### Stage 1: Candidate Extraction

Each unlabeled clinical message was processed by 3 complementary extraction methods operating in parallel:

1. An LLM-based extractor that reads the full message and proposes candidate text spans with domain hints for the 4 entity categories (Medication, Mentation, Mobility, and What Matters). The LLM was prompted (see Textbox A2) with domain definitions and labeling function specifications for each category and instructed to return short, specific (atomic) spans as exact substrings of the message, with a maximum of 30 spans per message.
2. A deterministic lexical candidate generator using regular expressions to identify known medication names, dosage patterns (eg, dose units, routes, frequencies), medication action keywords (eg, “discontinue,” “start,” “hold”), mentation keywords (eg, “confusion,” “agitation,” “oriented”), mobility keywords (eg, “ambulat,” “transfer,” “fall,” “wheelchair”), and What Matters keywords (eg, “prefers,” “refuses,” “hospice,” “DNR”). Disposition language co-occurring with family or next-of-kin mentions was also captured as What Matters candidates.
3. The gold-trained fine-tuned Bio-ClinicalBERT model from Stage 1 of the 4M-ER pipeline, which contributed additional candidate spans based on learned contextual patterns. A confidence threshold of 0.5 was applied to filter low-confidence predictions, and a maximum of 30 candidates per message was retained.

#### Stage 2: Adjudication

Candidate spans from Stage 1 were merged by proximity and domain, grouped, and submitted to an LLM adjudicator (see Textbox A3). For each span group, the adjudicator made a classification decision: whether the span constitutes a valid 4M entity and, if so, which domain it belongs to. The adjudicator was required to provide a verbatim evidence quote from the source text and a numerical confidence score (0 to 1) to support each decision. Spans classified as non-4M entities (is_4m=false) or assigned the label NO_LABEL were discarded. Evidence quotes that could not be verified as exact substrings of the original message were flagged and the confidence was capped at 0.6.

#### Stage 3: Skeptic Review

Every span that Stage 2 labeled as a 4M entity was subjected to a final adversarial review by a separate LLM call (the “skeptic”; see Textbox A4). The skeptic was instructed to challenge the proposed label and reject weak, ambiguous, or borderline annotations. Only spans that maintained high confidence through both adjudication (2: 0.75) and skeptic review (2: 0.80) were retained in the final silver dataset. Overlapping spans were resolved by retaining the higher-confidence span. This 3-stage extract-adjudicate-challenge design maximizes precision of the silver labels at the cost of some recall.

##### Silver Dataset Characteristics

The resulting silver dataset comprised 2,502 labeled messages, each containing at least 1 annotated entity span, with the following token-level distribution:

| **Token Label** | **Count** | **Percentage** |
| --- | --- | --- |
| O (outside) | 62,303 | 76.37% |
| Medication (B+I) | 13,992 | 72.6% of entities |
| What Matters (B+I) | 2,273 | 11.8% of entities |
| Mobility (B+I) | 1,907 | 9.9% of entities |
| Mentation (B+I) | 1,106 | 5.7% of entities |

The silver dataset exhibited a strong skew toward Medication entities (72.6% of entity tokens) compared with the gold training set (61.7%), with reduced representation of What Matters (11.8% vs 21.7%), Mobility (9.9% vs 9.5%), and Mentation (5.7% vs 7.2%).

This imbalance reflects the inherent strength of lexical and pattern-based extraction for pharmacological entities, which have distinctive surface forms (drug names, dosage patterns), relative to more linguistically diverse categories such as What Matters, where relevant spans tend to be conversational, indirect, and context-dependent.

##### Silver Data Filtering

To mitigate the distributional mismatch between the medication-heavy silver data and the more balanced gold evaluation distribution, the silver dataset was filtered to retain only messages where Medication entities comprised less than 50% of all entity tokens. This reduced the silver training set from 2,502 to 636 examples, enriching the remaining data for the underrepresented categories (Mentation, Mobility, and What Matters).

**Textbox A2.**
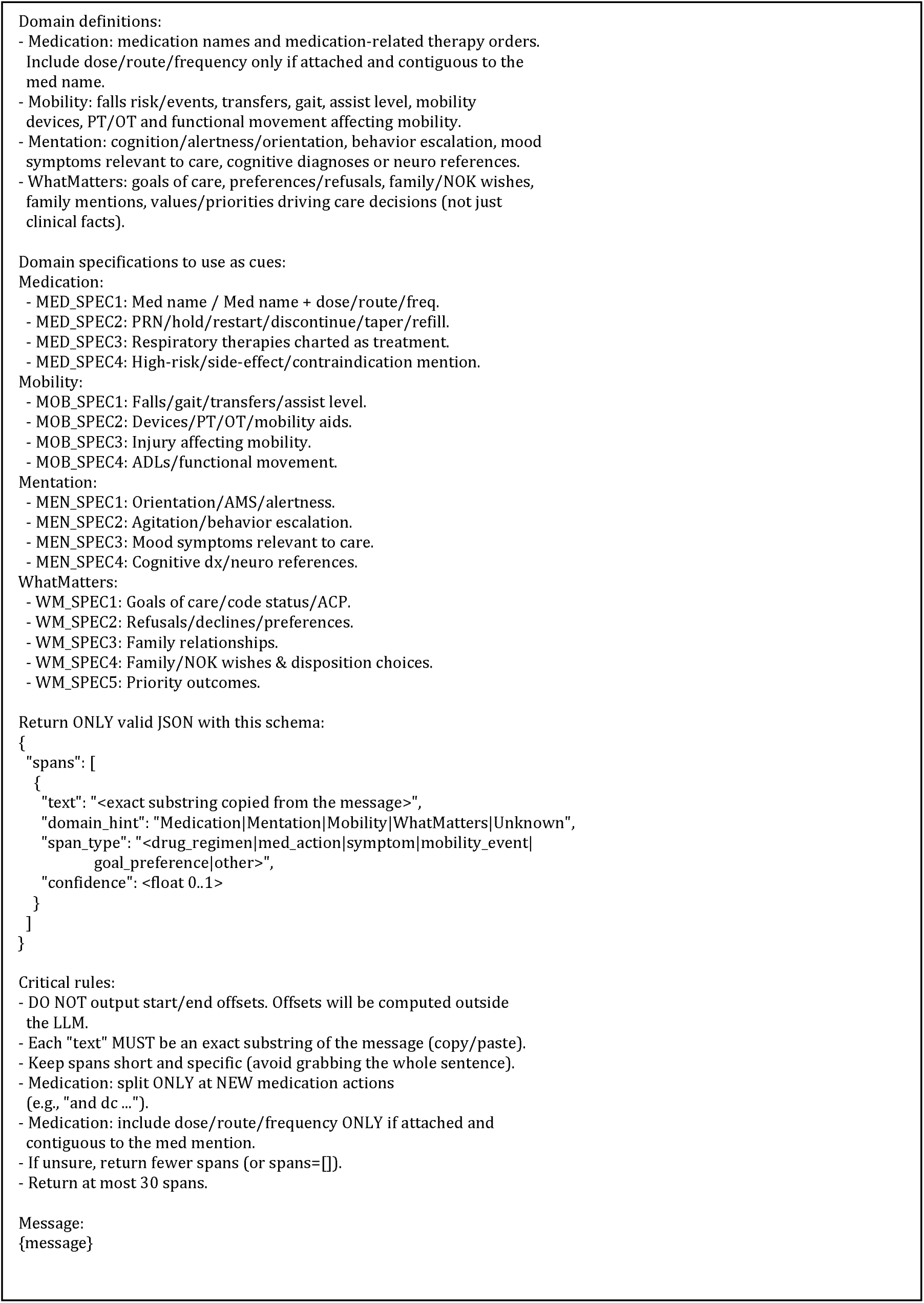
Stage 1 prompt for candidate span extraction in the silver data generation pipeline.

**Textbox A3.**
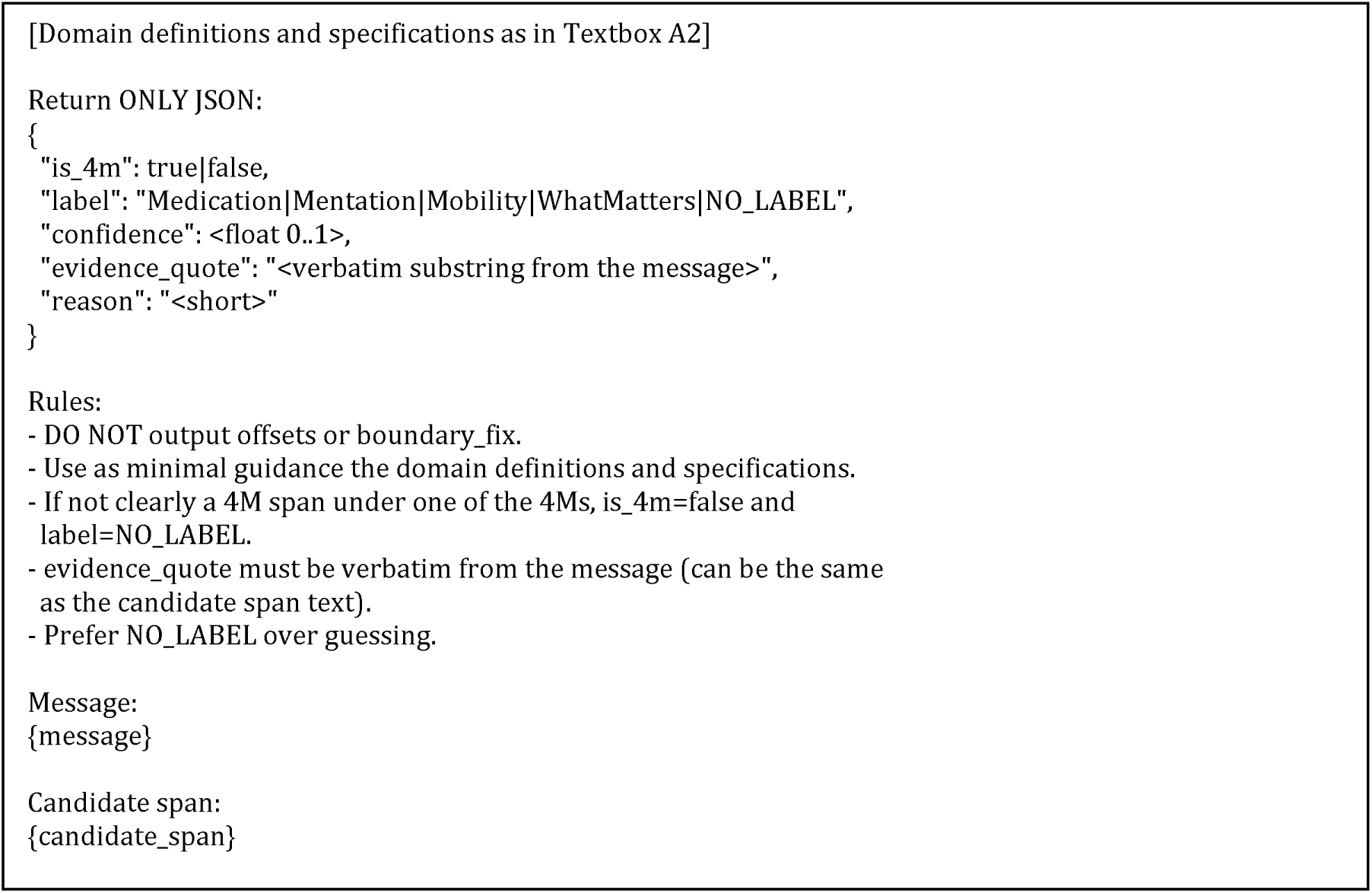
Stage 2 adjudication prompt for the silver data generation pipeline.

**Textbox A4.**
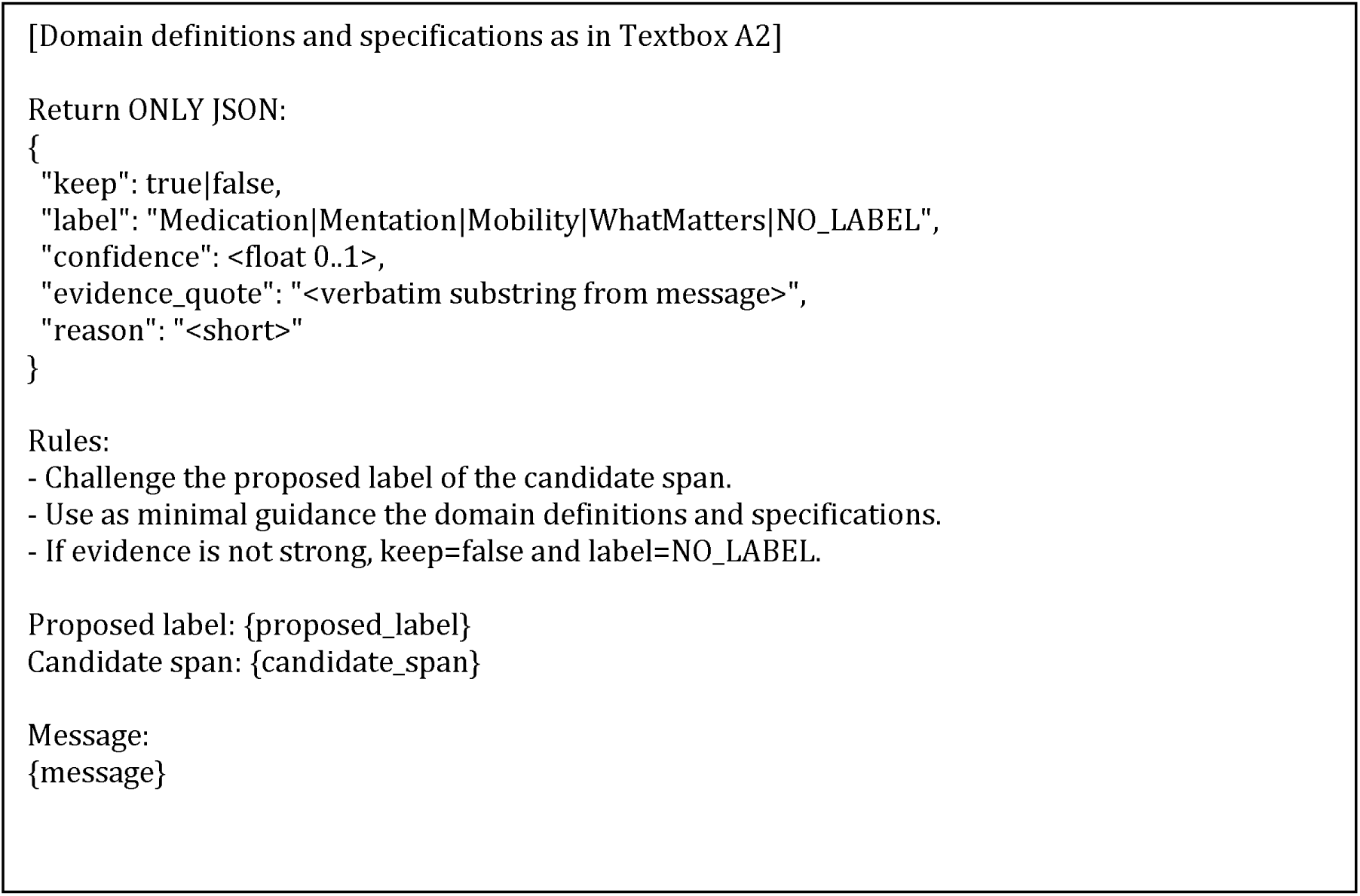
Stage 3 skeptic review prompt for the silver data generation pipeline.

